# Machine learning analysis reveals biomarkers for the detection of neurodegenerative diseases

**DOI:** 10.1101/2022.02.15.22270625

**Authors:** Simon Lam, Muhammad Arif, Xiya Song, Mathias Uhlen, Adil Mardinoglu

## Abstract

It is critical to identify biomarkers for neurodegenerative diseases (NDDs) to advance disease diagnosis and accelerate drug discovery for effective treatment of patients. In this work, we retrieved genotyping and clinical data from 1223 UK Biobank participants to identify genetic and clinical biomarkers for NDDs, including Alzheimer’s disease (AD), Parkinson’s disease (PD), motor neuron disease (MND), and myasthenia gravis (MG). Using a machine learning modelling approach and Monte Carlo randomisation, we identified 16 informative clinical variables for predicting AD, PD, MND, and MG. In a multinomial model, these clinical variables could correctly predict the diagnosis of one of the four diseases with an accuracy of 88.3%. In addition to clinical biomarkers, we also explored genetic biomarkers. In a genome-wide association study of AD, PD, MND, and MG patients, we identified single nucleotide polymorphisms (SNPs) implicated in several craniofacial disorders such as apnoea and branchiootic syndrome. We found evidence for shared genetic risk loci across NDDs, including SNPs in cancer-related genes and SNPs known to be associated with non-brain cancers such as Wilms tumour, leukaemia, and pancreatic cancer. Our analysis supports current knowledge regarding the ageing-related degeneration/cancer shift.

**Significance statement:** This study highlights the potential for hypothesis-free mathematical modelling of easily measured clinical variables to identify diagnostic biomarkers for neurodegenerative diseases (NDDs). Prior to this study, the focus in NDD research has surrounded toxic species such as amyloid beta and α-synuclein, but this approach has not enjoyed success at clinical trial. Here, we studied Alzheimer’s disease, Parkinson’s disease, motor neuron disease, and myasthenia gravis by constructing and inspecting a multinomial based on demographics and blood and urine biochemistry. Cognitive measures were important for the predictive power of the model. Model weights correctly indicated multiple trends reported in the literature. Separately, genome-wide association indicated a shared risk profile between NDD and cancer, which has also been reported in the literature.

## Introduction

Neurodegenerative diseases (NDDs) pose a significant public health problem due to ageing worldwide. The global burden of NDDs is increasing (Feigin et al., 2021; Nichols et al., 2019; Ray Dorsey et al., 2018). Alzheimer’s disease (AD) is characterised by the accumulation of amyloid-beta plaques and tau protein neurofibrillary tangles. These toxic species result in the destruction of cholinergic neurons and cause cognitive decline, leading to dementia. Parkinson’s disease (PD) is molecularly characterised by α-synuclein inclusions in dopaminergic neurons in the substantia nigra, resulting in dopamine depletion and movement disorders. Motor neuron disease (MND) and myasthenia gravis (MG) are movement disorders that affect motor neurons and muscle, respectively, with MG also being an autoimmune disease. Individuals with PD or MND may also develop dementia.

Despite decades of research and clinical trials, there are currently no treatments to reverse the damage done by neurodegeneration. Current research has focused on toxic species such as amyloid-beta and tau in AD and α-synuclein in PD. Still, treatments derived from the research have been largely unsuccessful at clinical trials (Lam et al., 2020). Alternative pharmacological and nonpharamacological targets need to be identified to enable early diagnosis and accelerate drug discovery to treat patients with NDDs effectively, and there is already great interest in these fields (Bayraktar et al., 2021; Lam et al., 2021; Zool et al., 2020).

In this study, we retrieved genotyping and clinical data from 1223 participants in the UK Biobank (Sudlow et al., 2015), identified genetic predispositions to NDDs and discovered diagnostic markers. We used clinical data to construct a predictive multinomial general linear model for AD, PD, MND, and MG with 88.3% accuracy, using Monto Carlo randomisation to select the clinical variables included in the model. In addition to the high valid positive rate, the model weights correctly identified trends among disease patients concerning blood and urine biochemistry and cognitive function. We, therefore, propose this data-driven machine learning method as an ideal first step for identifying new biomarkers for detection of the diseases. We also used genotype data to identify genetic predispositions to neurodegeneration. Our results indicated that many risk alleles are shared across multiple NDDs and are also shared with non-brain cancers. Given the understanding of a degeneration/cancer shift with ageing, our results indicated that the shift may be screened at the genetic level.

## Results

### Blood and urine biochemistry and cognitive trends inform NDD status

We retrieved clinical data from 1223 patients with AD, PD, MND, or MG, of which 1072 also had Affymetrix Axiom genotyping data on 820967 SNPs, from the UK Biobank (Table 1). The mean age of participants with AD, PD, MND, and MG was 62.8, 62.1, 59.4, and 60.2, respectively, compared with 53.7 for control participants. The proportions of each group who were male were 56.6%, 63.3%, 69.2%, and 46.6%, respectively, compared to 45.3% male for control participants. The cohort was made up of 89.5%, 91.3%, 90.8%, and 91.4% British participants, respectively, compared with 87.1% British for control participants.

**Table 1.** Characteristics of UK Biobank NDD dataset

| Disease | Total | With<br>genotyping<br>data |
| --- | --- | --- |
| AD | 152 | 129 |
| PD | 948 | 832 |
| MND | 65 | 60 |
| MG | 58 | 51 |
| Control | 116559 | 446 |
| Total<br>(excluding<br>controls) | 1223 | 1072 |

Patients with NDDs exhibited changes to blood and urine biochemical markers (Figure 1, Supplementary Table 1). Participants with AD had significantly higher cystatin C levels. PD participants had lower alanine aminotransferase (ALT), albumin, apolipoprotein A, calcium, cholesterol, LDL, and phosphate, but elevated sodium and testosterone, compared to control. Participants with MND had significantly higher ALT, cystatin C, and testosterone compared to control. MG patients had decreased albumin, cholesterol, and LDL, but increased cystatin C compared to control. All NDD patients took longer to correctly identify matches in cognitive testing (Supplementary Table 2), compared to control. In the prospective memory test, AD patients were the most likely not to recall the instruction after two attempts and showed the least improvement between first and second visits.

**Figure 1.**
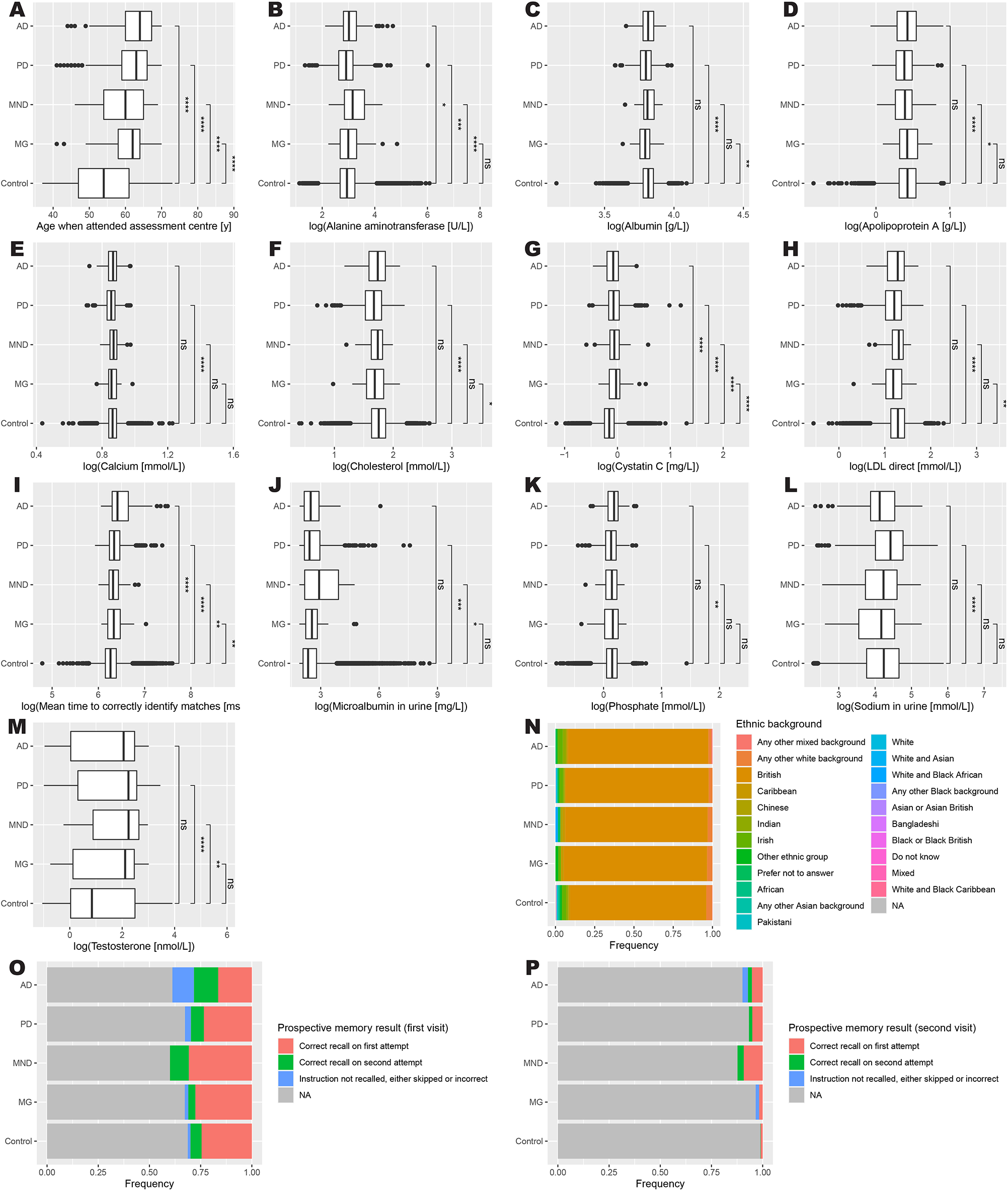
Distribution of clinical measures by diagnosis. A) Age when attended assessment centre. B) Alanine aminotransferase. C) Albumin. D) Apolipoprotein A. E) Calcium. F) Cholesterol. G) Cystatin C. H) LDL direct. I) Mean time to correctly identify matches. J) Microalbumin in urine. K) Phosphate. L) Sodium in urine. M) Testosterone. N) Ethnic background. O) Prospective memory result (first visit). P) Prospective memory result (second visit). Data represent all participants, including those whose samples which were not used in the training or testing of the multinomial model. AD, Alzheimer’s disease (n = 152); PD, Parkinson’s disease (n = 948); MND, motor neuron disease (n = 65); MG, myasthenia gravis (n = 58); Control (n = 116559). Mean comparisons were performed using t-tests. *, p ≤ 0.05; **, p ≤ 0.01; ***, p ≤ 0.001; ****, p ≤ 0.0001; ns, p > 0.05.

From these results, we clearly see that there are trends in biochemical and cognitive measures, which seem to differ in direction between NDDs. We therefore sought next to generate a predictive model based on these results.

### A multinomial model classified neurological diseases and identified relevant biomarkers

We generated a multinomial generalised linear model to predict AD, PD, MND, MG, and control in a single test. We selected the clinical measures with good coverage among participants to create this model. We used a Monte Carlo randomised sampling method to optimise the combined measures included in the model (see Methods). The confounders in our final model can be summarised as follows: demographics (age, ethnicity), blood test measures (ALT, albumin, apolipoprotein A, calcium, cholesterol, cystatin C, LDL, phosphate, testosterone), urine test measures (microalbumin, sodium), and cognitive test measures (prospective memory test, reaction test). The multinomial model had a true positive rate of 88.3% on unseen data (Table 2 & Supplementary Table 3).

**Table 2.**
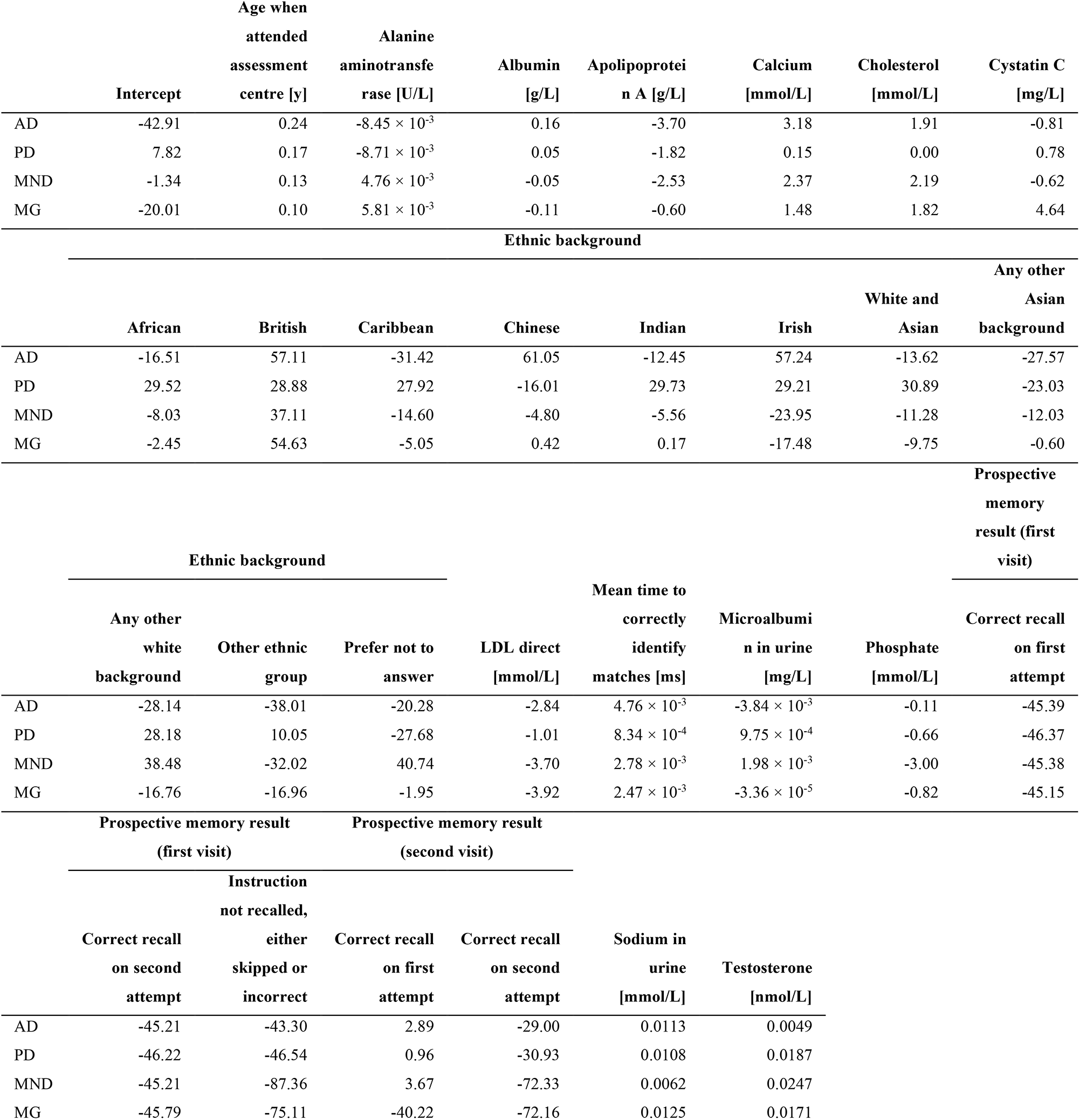
Coefficients of the multinomial generalised linear model.

We performed leave-one-out cross validation to demonstrate the essentiality of each of the variables included in the model in terms of its contribution to predictive power (Supplementary Table 4). We constructed and tested new multinomial models after singly, separately, omitting each variable. We found that the model was robust to single variable omission, with most models performing at around 87%-88% accuracy. The biggest drops arose from omission of prospective memory result (first visit) (76.0% accuracy), age (85.8% accuracy), and testosterone (87.0%). Omission of LDL, cystatin C, mean time to correctly identify matches, sodium, and apolipoprotein A all produced models with 87.7% accuracy. Age and cognition appeared to be the most important factors contributing to model accuracy.

We inspected the coefficients assigned to each of the clinical variables included in the model. AD was more likely to be predicted for patients with low ALT, apolipoprotein A, cystatin C, LDL, and urine microalbumin. High calcium, cholesterol, and urine sodium were also characteristics of AD identified by the model. AD was the disease most tightly linked to advanced age, and patients of British, Chinese, or Irish background, were more likely to be predicted to have AD by the model, as did those who took the longest to identify matches. PD was assigned to those patients with advanced age, low ALT, low apolipoprotein A, and high testosterone. People of Chinese background or any other Asian background were less likely to be predicted to have PD. The model assigned MND to those patients with high ALT calcium, cholesterol, and testosterone, but low apolipoprotein A and phosphate. Patients who answered the ethnicity question with “any other white background” were more likely to be assigned MND by the model. The model linked MG with high ALT and cystatin C, but low albumin and LDL. Patients of British background were more likely to be assigned MG, whereas those of Irish and those who answered “any other white background” were less likely.

Across all diseases, the model applied a negative coefficient for the prospective memory test at the first visit, regardless of the result. Patients who took more attempts to recall correctly, or did not recall at all, were more likely to be assigned AD by the model than any other disease. Interestingly, the coefficients for PD were very similar regardless of the result. Together with the small coefficient given to the match identification test, the model suggests that cognitive measures were not very important for the classification of PD. In contrast, patients not recalling the instruction were given very negative coefficients for MND and MG, indicating that these patients are more likely to have AD or PD instead.

Interestingly, at the second visit, the coefficients were always lower if the patient took two attempts rather than one, which appears counter-intuitive for AD. However, AD also had the least negative coefficient, indicating that patients who took two attempts to recall the instruction at the second visit were more likely to be assigned AD.

The model also identified numerous interactions with blood and urine biochemical levels, indicating elevated ALT in MND and MG, but decreased in AD and PD; sharply decreased apolipoprotein A in AD and MND, but more modest decreases in PD and MG; increases in calcium and cholesterol in AD, MND, and MG, but changes in these molecules not being significant in PD; decreases in LDL and phosphate in any disease, but most strikingly in MG and MND, respectively; and increases in sodium and testosterone in all diseases. That the model was able to suggest these directional changes, which could not be so clearly concluded by t-tests alone (Figure 1), indicates the potential for machine learning techniques to identify possible biomarkers for classification in the absence of an understanding by the model of the underlying biology.

### Genome-wide association uncovered shared heritable factors between neurological diseases

In the multinomial model, we identified a heritable variable, ethnicity, to predict neurodegeneration. Therefore, we hypothesised that there might be more heritable factors that could alter a person’s propensity to neurodegenerative disease. To identify heritable factors, we analysed genotyping data originating from an Affymetrix Axiom array, which covers 836,727 single-nucleotide polymorphisms (SNPs) (Figure 2, Supplementary Figure 3, Supplementary Table 5). We found significant SNPs which are also associated with brain and non-brain diseases. Interestingly, we found SNPs associated with the same diseases across the different patient groups. For example, AD and MG patients both had SNPs associated with apnoea, branchiootic syndrome, and asthma; AD and PD patients both had SNPs associated with PD and cognitive function; and all patients had SNPs associated with non-brain cancers. One of these non-brain cancers was Wilms tumour, a type of kidney cancer, for which an intronic SNP in the VPS41 gene was shared across all four patient groups. Other non-brain cancers included leukaemia, pancreatic cancer, oesophageal carcinoma, and colorectal cancer.

**Figure 2.**
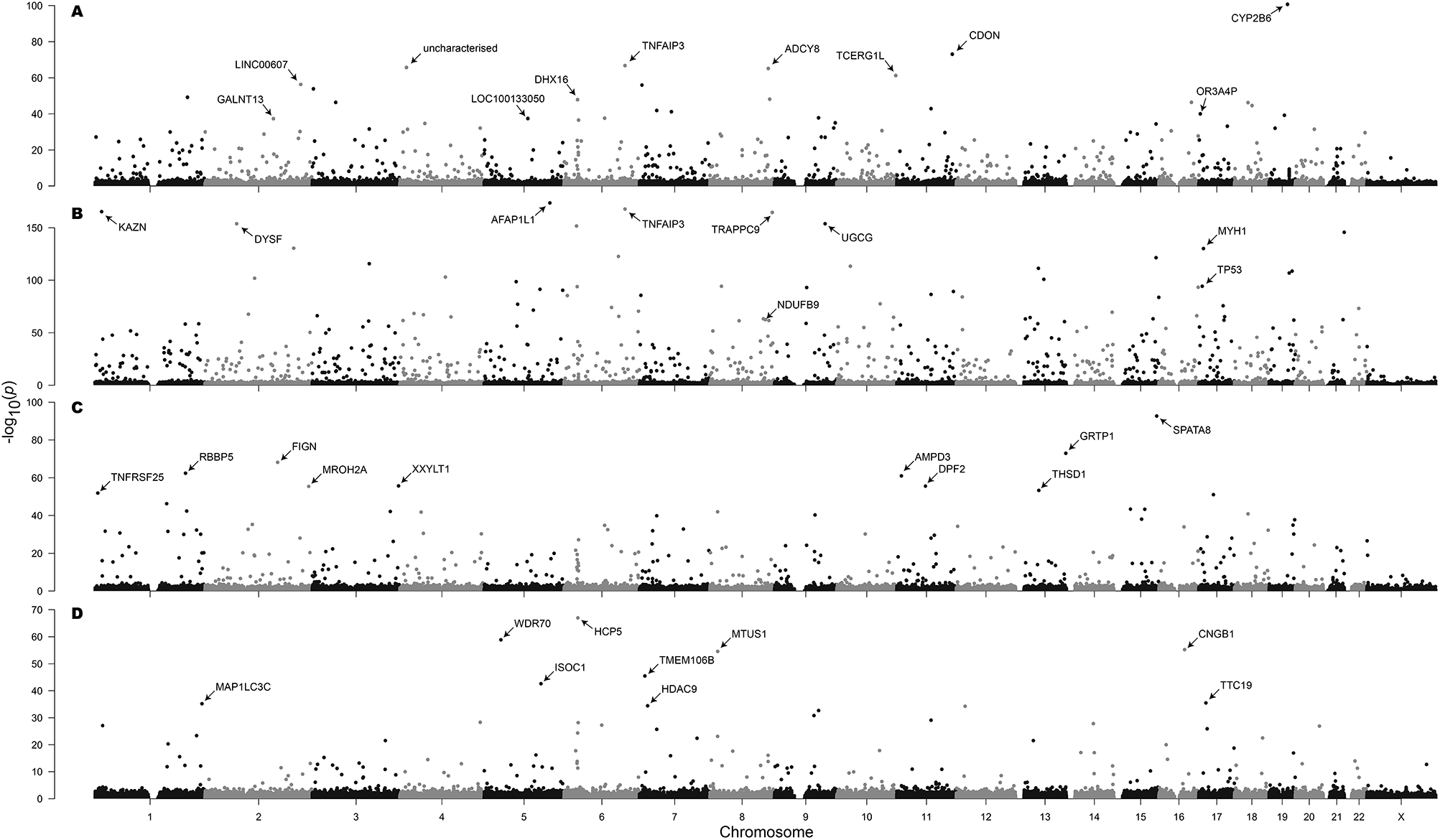
Genome-wide association with A) Alzheimer’s disease, B) Parkinson’s disease, C) motor neuron disease, and D) myasthenia gravis in the Affymetrix Axiom Biobank Array. Selected SNPs are labelled with the symbol of the containing or nearest gene.

Considering the genes affected by the SNPs we identified, we found a vast number of significant SNPs within genes associated with cancer, its hallmarks, or neurotransmission. An intronic SNP in the scaffold protein interactor GAB2 was associated with all four diseases. A missense mutation in the TNFα response gene TNFAIP3 gene was highly associated with AD and PD, but not MND or MG. AD was also associated with SNPs in the cytochrome P450 gene CYP2B6, adenylate cyclase ADCY8, and signalling gene PIK3C3 (PI3K). PD was highly associated with SNPs in dysferlin (DYSF), TP53, and numerous cytoskeleton genes such as AFAP1L1 and MYH1. MND was associated with SNPs in retinoblastoma protein interactor RBBP5, tumour necrosis factor family member TNFRSF25, and several cytoskeleton function genes, including tropomyosin TPM1 and troponin TNNI3. MND was also associated with SNPs in spermatogenesis associated genes such as SPATA8 and SPAG16. MG was associated with SNPs in hallmarks of cancer genes such as the microtubule-associated tumour suppressor MTUS1, leukocyte-associated gene LAIR2, and Cbl proto-oncogene CBLB. MG was also associated with SNPs in the brain-and neuron-specific genes, including cerebellin 4 precursor (CBLN4), potassium channel KCNH5, adenylate cyclase 8 (ADCY8), and autism susceptibility candidate gene AUTS2.

These results indicated that the same SNPs may be associated with susceptibility to more than one NDD. Further, with the appearance of SNPs in genes related to cancer and hallmarks of cancer, there may be SNPs associated with both NDD and cancer. Since neurodegeneration and cancer are antagonistic, that is, patients with neurodegeneration are less likely to develop cancer, and vice versa, the appearance of cancer-related SNPs is consistent.

## Discussion

This study shows the value of big biological data in driving hypothesis-free studies towards the early diagnosis of NDDs. We first constructed a multinomial model that could predict AD, PD, MND, or MG with an accuracy of 88.3%. The clinical variables that we used to generate the model were selected by a Monte Carlo randomisation method, and the coefficients fit the model closely reflect existing knowledge.

One clinical variable included in the model was ethnicity. Therefore, we next carried out a genome-wide association study to identify more heritable factors which might predispose an individual to AD, PD, MND, or MG. While we could not account for the variability in the data due to ethnicity, we found commonalities across all four diseases – particularly the appearance of SNPs associated with non-brain cancers. When training our multinomial model, the theoretical search space was 2^40^. Had we included all clinical measures; the search space would have exceeded 2^400^. We were able to identify biologically significant biomarkers through Monte Carlo exploration of a small fraction of the search space shows the potential for such untargeted approaches to biomarker discovery.

Identifying easily measurable clinical variables by simple blood and urine biochemical tests is favourable as these collection methods are minimally invasive and widely accessible. In addition to these, our model also identified age, ethnicity, and cognitive function as confounders for neurological disease. Interestingly, our model did not include sex, although it did identify testosterone.

Our model assigned coefficients to the clinical variables for each disease. These coefficients are weights used by the model to aid classification and can be positive, negative, or zero. They can be interpreted as predictions into the importance and impact of each clinical variable on a person’s susceptibility to each disease. For instance, the model assigned a negative coefficient to ALT for AD and PD, but a positive coefficient for MND and MG. This would suggest low ALT being associated with the former disorders and high ALT being associated with the latter diseases.

In the case of ALT, this association of low ALT with AD is consistent with the literature (Lu et al., 2021). Other brain diseases such as intracerebral haemorrhage have been linked with elevated ALT (Nho et al., 2019). In addition, our model correctly suggested low apolipoprotein A in all four diseases, which is consistent with the literature for AD (Zuin et al., 2021), PD (Qiang et al., 2013), and MND (Mariosa et al., 2017). The positive coefficients for calcium in AD and PD are consistent with evidence from familial AD studies (Ryan et al., 2020) and α-synuclein in mitochondria (Angelova et al., 2016; Zakharov et al., 2007). Elevated calcium in MND is also supported by studies on ANXA11 mutations that is associated with MND and increased basal calcium concentration (Nahm et al., 2020) and C9ORF72 (Dafinca et al., 2020). Diminished LDL in MG might be supported by Lrp4, which binds LDL, is an autoimmune target in MG (Koneczny & Herbst, 2019). Less favourable scores on cognitive tests (i.e., longer mean time to correctly identify matches, longer or no recall on a prospective memory test) appeared to be predictive for AD, which would be as expected due to its known association with cognitive decline. Finally, AD being given the lowest coefficient for testosterone is in keeping with the knowledge that AD is more prevalent in women and that low plasma testosterone is associated with AD in older adults.

There were some limitations to the model. Firstly, it could not capture all of the directional effects of each clinical variable on each disease. An example of this was the significant negative coefficient for LDL in MND, suggesting that MND patients might have lower serum LDL, whereas this is not the case according to the literature(Chen et al., 2018; Zeng & Zhou, 2019). Secondly, although the model had an accuracy of 88.3%, it was better at predicting some diseases than others. Both of these limitations were due to the relatively low numbers of patients with AD, MND, and MG. We found that the model was best at predicting PD and control and performed less well when predicting the other diseases. Because of this, we indicate that the model may be improved with the analysis of more samples and more consistent data collection across all patients. This limitation, however, did not detract from the model’s usefulness to be deconstructed to predict biomarkers, and therefore all four diseases were included in the model.

We identified heritable factors to add into the multinomial model after identifying ethnicity as a predictive factor for neurological diseases. Although we could not improve the model using SNP data, we instead found many SNPs that were significantly linked with multiple neurological diseases and diverse cancer types. The appearance of cancer-related SNPs is significant and is consistent with the degeneration/cancer antagonism, which is well reported in the literature (Aramillo Irizar et al., 2018). Our current work suggests that the genetic risk factors for neurological diseases are also risk factors for cancer. This means that patients might initially be susceptible to both neurodegeneration and cancer before exhibiting an antagonistic shift with age. The co-occurrence of neurodegenerative and cancer SNPs could have arisen due to a selective pressure to bolster the cellular senescence response, a tumour-suppressing process which also contributes to both degeneration and cancer when used in an aberrant manner (Campisi, 2013). This would be consistent with our observation of SNPs within TNF family member genes, as these genes often have roles associated with cell turnover. However, since we did not assess SNPs in patients with a cancer diagnosis, this result should not be over-interpreted.

Taken together, the identification of known blood and urine biomarkers by a machine learning model with no knowledge of the underlying biology, and confirmation of a link between NDDs and cancer are promising for the acceleration of the first step towards biomarker discovery.

## Methods

### Acquisition of data

Data were obtained from UK Biobank (Sudlow et al., 2015). Samples were accepted as being Alzheimer’s disease (AD), myasthenia gravis (MG), motor neuron disease (MND), or Parkinson’s disease (PD) if any of the respective diseases were recorded in the data, even if other conditions were also recorded. Control samples were accepted as those without any recorded diseases (Table 1).

### Generation and leave-one-out cross validation of the multinomial model

To generate the multinomial model, clinical data including demographics, sight and hearing problems, diabetes diagnosis, stroke diagnosis, medication and treatment, illness, operations, cognitive and mental measures, brain measurements, blood and urine tests, and adverse events and death were obtained from UK Biobank. Clinical variables were removed if less than 75% of samples had a value recorded for that variable. Categorical measures with only one category were also removed. After quality control, 40 clinical variables were considered.

Monte Carlo randomisation was used to randomly sample clinical variables as independent variables in a multinomial model. Samples with any missing values across the selected clinical variables were dropped. The multinomial model was constructed using the R nnet package (version 7.3-14, https://cran.r-project.org/web/packages/nnet/index.html, accessed 2021-06-01). The dependent variable was disease. The dataset was randomly split into training (70%) and test sets (30%) to assess the accuracy (true positive rate) of the model on test set data. The model with the highest accuracy after 1000 random samplings was accepted for further consideration.

To find the model with the local maximum accuracy, clinical variables not in the model were added, and clinical variables already in the model were removed, individually. If, after each model change, the model’s accuracy improved, then that change was kept; if otherwise, then that change was reverted.

To perform leave-one-out cross validation on the multinomial model, variables were singly removed from the model, separately, and multinomial models were constructed and tested, as above.

### Genotyping analysis

Raw genome-wide genotyping data were obtained from the UK Biobank. These data had been generated from the UK Biobank Axiom Array. Axiom Analysis Suite (version 5.1.1, ThermoFisher, accessed 2021-06-01) was used to perform quality control and analysis on the raw data to determine patient genotypes at each single nucleotide polymorphism (SNP) position, using the Best Practices Workflow. Allele frequencies were computed from genotype data for each disease class. To detect significant differences in allele frequencies between disease and control, Cochran-Armitage trend tests were performed assuming a codominant allele model. The test statistics, exact p-values, and asymptotic p-values were recorded. CATTexact (version 0.1.1, https://cran.r-project.org/web/packages/CATTexact/index.html, accessed 2021-06-01) R package was used for the computation of Cochran-Armitage p-values(Mehta et al., 1992). qqman (version 0.1.8, https://cran.r-project.org/web/packages/qqman/index.html, accessed 2021-06-01) R package was used to plot the Manhattan and quantile-quantile plots. All bioinformatic and statistical analyses were performed in R (version 4.0.2) unless otherwise indicated.

## Data Availability

All data produced are available online at https://github.com/SimonLammmm/ukbb-ndd-ml

## Author contributions

M.U. and A.M. supervised the study. S.L. designed the study, performed all the analyses, and analysed all the results. S.L. wrote the manuscript with input from M.A., X.S., M.U., and A.M.

## Acknowledgements

This research has been conducted using the UK Biobank Resource under Application Number 64488.

The authors acknowledge use of the research computing facility at King’s College London, *Rosalind* (https://rosalind.kcl.ac.uk, accessed 2021-06-01).

This work was financially supported by Knut and Alice Wallenberg Foundation (grant number 2017.0303) to A.M.

## SI figure legends

**Supplementary Figure 1.**
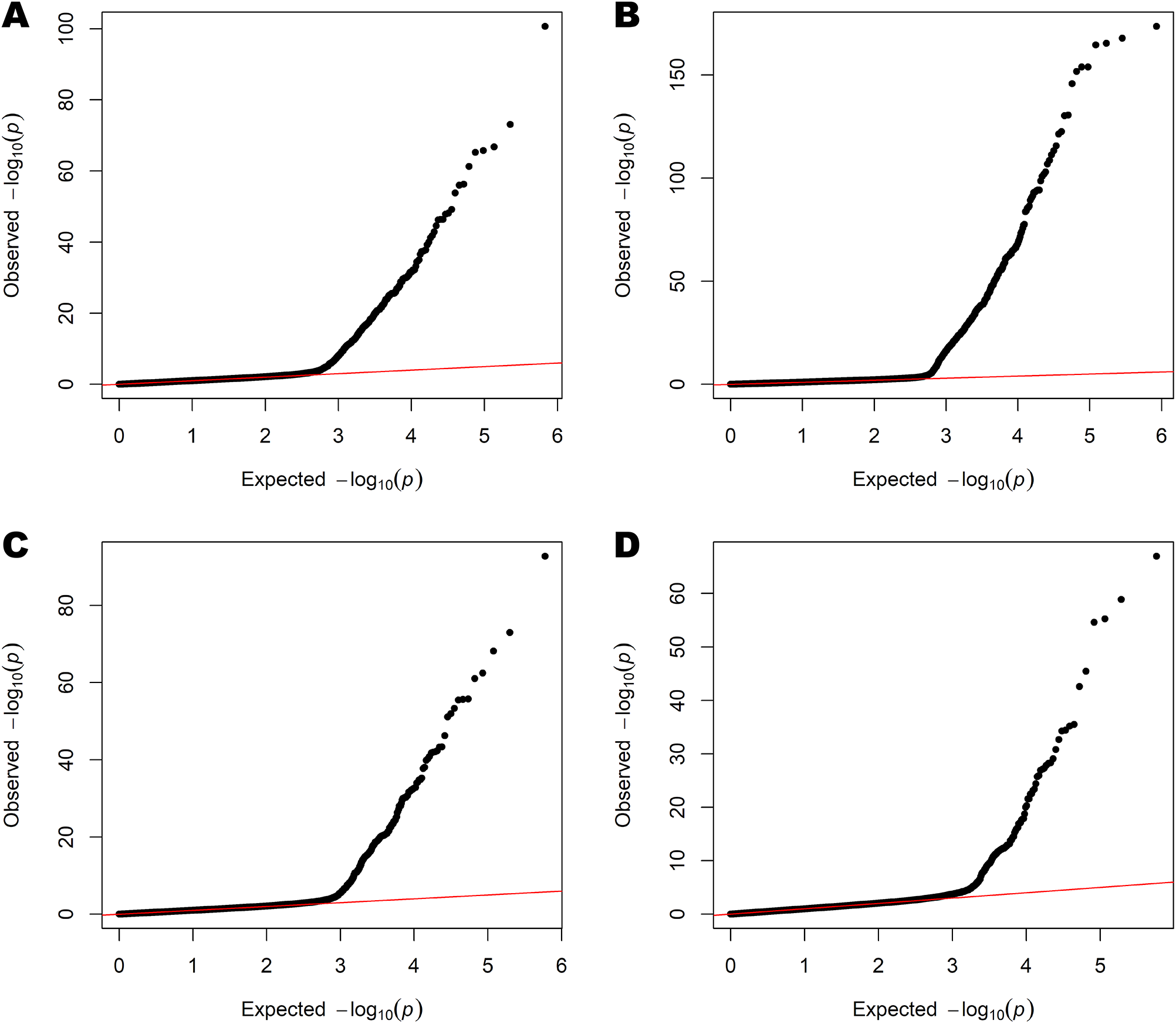
Normal quantile-quantile plots of A) Alzheimer’s disease, B) Parkinson’s disease, C) motor neuron disease, and D) myasthenia gravis SNPs in the Affymetrix Axiom Biobank Array. All p-values are asymptotic Cochran-Armitage trend test p-values. *Except for age, t-tests were performed after log transformation

**Supplementary Table 1.** Summary statistics of UK Biobank NDD dataset

| Clinical marker | Disease | Mean | 95% CI |  | p-value (t-test)* |
| --- | --- | --- | --- | --- | --- |
| **Age when attended**<br>**assessment centre [y]** | AD | 62.77 | 61.86 | 63.68 | $2.17 \times 10^{-43}$ |
| | PD | 62.13 | 61.79 | 62.48 | $1.80 \times 10^{-255}$ |
| | MND | 59.40 | 57.78 | 61.02 | $1.92 \times 10^{-9}$ |
| | MG | 60.17 | 58.50 | 61.84 | $2.12 \times 10^{-10}$ |
|  | Control | 53.74 | 53.69 | 53.78 |  |
| **Alanine**<br>**aminotransferase**<br>**[U/L]** | AD | 24.06 | 21.81 | 26.31 | 0.034 |
| | PD | 20.81 | 19.75 | 21.87 | $1.20 \times 10^{-5}$ |
| | MND | 28.21 | 24.80 | 31.62 | $1.82 \times 10^{-4}$ |
|  | MG | 24.59 | 19.68 | 29.50 | 0.402 |
|  | Control | 22.07 | 21.99 | 22.14 |  |
| **Albumin [g/L]** | AD | 45.30 | 44.87 | 45.74 | 0.571 |
| | PD | 44.64 | 44.47 | 44.81 | $8.21 \times 10^{-17}$ |
|  | MND | 45.40 | 44.81 | 45.99 | 0.951 |
| | MG | 44.41 | 43.74 | 45.07 | $5.61 \times 10^{-3}$ |
|  | Control | 45.43 | 45.42 | 45.45 |  |
| **Apolipoprotein A**<br>**[g/L]** | AD | 1.55 | 1.50 | 1.59 | 0.531 |
| | PD | 1.48 | 1.47 | 1.50 | $4.53 \times 10^{-16}$ |
|  | MND | 1.47 | 1.41 | 1.53 | 0.0144 |
|  | MG | 1.58 | 1.51 | 1.65 | 0.590 |
|  | Control | 1.56 | 1.56 | 1.56 |  |
| **Calcium [mmol/L]** | AD | 2.38 | 2.36 | 2.39 | 0.977 |
| | PD | 2.35 | 2.35 | 2.36 | $1.21 \times 10^{-13}$ |
|  | MND | 2.39 | 2.37 | 2.42 | 0.225 |
|  | MG | 2.36 | 2.34 | 2.39 | 0.255 |
|  | Control | 2.38 | 2.38 | 2.38 |  |
| **Cholesterol [mmol/L]** | AD | 5.70 | 5.52 | 5.88 | 0.0814 |
| | PD | 5.37 | 5.30 | 5.44 | $2.92 \times 10^{-33}$ |
|  | MND | 5.64 | 5.42 | 5.85 | 0.115 |
|  | MG | 5.48 | 5.17 | 5.80 | 0.0203 |
|  | Control | 5.85 | 5.84 | 5.86 |  |
| **Cystatin C [mg/L]** | AD | 0.94 | 0.91 | 0.96 | $1.03 \times 10^{-7}$ |
| | PD | 0.96 | 0.94 | 0.97 | $8.85 \times 10^{-52}$ |
| | MND | 0.96 | 0.91 | 1.00 | $3.22 \times 10^{-4}$ |
| | MG | 0.99 | 0.94 | 1.04 | $2.92 \times 10^{-6}$ |
|  | Control | 0.87 | 0.87 | 0.87 |  |
| **LDL direct [mmol/L]** | AD | 3.57 | 3.43 | 3.71 | 0.125 |
| | PD | 3.36 | 3.30 | 3.41 | $1.50 \times 10^{-25}$ |
|  | MND | 3.57 | 3.41 | 3.72 | 0.399 |
| | MG | 3.34 | 3.12 | 3.57 | $6.54 \times 10^{-3}$ |
|  | Control | 3.67 | 3.66 | 3.67 |  |
| **Mean time to**<br>**correctly identify**<br>**matches [ms]** | AD | 693.30 | 654.25 | 732.35 | $5.15 \times 10^{-14}$ |
| | PD | 591.66 | 583.36 | 599.96 | $2.99 \times 10^{-34}$ |
| | MND | 579.71 | 551.68 | 607.74 | $6.22 \times 10^{-3}$ |
| | MG | 588.43 | 555.41 | 621.45 | $3.54 \times 10^{-3}$ |
|  | Control | 544.18 | 543.55 | 544.81 |  |
| **Microalbumin in urine [mg/L]** | AD | 24.03 | 15.01 | 33.05 | 0.212 |
| | PD | 28.70 | 21.01 | 36.39 | $4.04 \times 10^{-3}$ |
|  | MND | 31.02 | 23.76 | 38.28 | 0.0314 |
|  | MG | 23.48 | 15.09 | 31.87 | 0.261 |
|  | Control | 20.92 | 20.47 | 21.38 |  |
| **Phosphate [mmol/L]** | AD | 1.18 | 1.16 | 1.21 | 0.106 |
| | PD | 1.14 | 1.13 | 1.15 | $7.17 \times 10^{-3}$ |
|  | MND | 1.14 | 1.11 | 1.18 | 0.566 |
|  | MG | 1.15 | 1.10 | 1.20 | 0.619 |
|  | Control | 1.16 | 1.16 | 1.16 |  |
| **Sodium in urine [mmol/L]** | AD | 74.39 | 67.77 | 81.02 | 0.447 |
| | PD | 89.43 | 86.57 | 92.29 | $6.70 \times 10^{-19}$ |
|  | MND | 74.53 | 64.05 | 85.01 | 0.562 |
|  | MG | 75.38 | 63.24 | 87.51 | 0.466 |
|  | Control | 78.48 | 78.22 | 78.74 |  |
| **Testosterone [nmol/L]** | AD | 7.19 | 6.29 | 8.10 | 0.114 |
| | PD | 8.47 | 8.08 | 8.85 | $1.86 \times 10^{-20}$ |
| | MND | 8.83 | 7.38 | 10.28 | $1.53 \times 10^{-4}$ |
|  | MG | 7.63 | 6.05 | 9.21 | 0.174 |
|  | Control | 6.66 | 6.62 | 6.70 | 0.0304 |

\*Except for age, t-tests were performed after log transformation

**Supplementary Table 2.** Summary statistics of UK Biobank NDD dataset

|  | AD |  | PD |  | MND |  | MG |  | Control |  |
| --- | --- | --- | --- | --- | --- | --- | --- | --- | --- | --- |
|  | Freq | % | Freq | % | Freq | % | Freq | % | Freq | % |
| **Ethnic background** |  |  |  |  |  |  |  |  |  |  |
| African | 0 | 0.00 | 2 | 0.21 | 0 | 0.00 | 0 | 0.00 | 984 | 0.85 |
| Any other Asian background | 0 | 0.00 | 3 | 0.32 | 0 | 0.00 | 0 | 0.00 | 479 | 0.41 |
| Any other Black background | 0 | 0.00 | 0 | 0.00 | 0 | 0.00 | 0 | 0.00 | 22 | 0.02 |
| Any other mixed background | 1 | 0.66 | 0 | 0.00 | 0 | 0.00 | 0 | 0.00 | 234 | 0.20 |
| Any other white background | 3 | 1.97 | 23 | 2.43 | 2 | 3.08 | 2 | 3.45 | 4390 | 3.78 |
| Asian or Asian British | 0 | 0.00 | 0 | 0.00 | 0 | 0.00 | 0 | 0.00 | 9 | 0.01 |
| Bangladeshi | 0 | 0.00 | 0 | 0.00 | 0 | 0.00 | 0 | 0.00 | 53 | 0.05 |
| Black or Black British | 0 | 0.00 | 0 | 0.00 | 0 | 0.00 | 0 | 0.00 | 5 | 0.00 |
| British | 136 | 89.47 | 864 | 91.33 | 59 | 90.77 | 53 | 91.38 | 101087 | 87.10 |
| Caribbean | 1 | 0.66 | 5 | 0.53 | 1 | 1.54 | 1 | 1.72 | 1018 | 0.88 |
| Chinese | 1 | 0.66 | 2 | 0.21 | 1 | 1.54 | 0 | 0.00 | 566 | 0.49 |
| Do not know | 0 | 0.00 | 0 | 0.00 | 0 | 0.00 | 0 | 0.00 | 55 | 0.05 |
| Indian | 3 | 1.97 | 8 | 0.85 | 0 | 0.00 | 0 | 0.00 | 1408 | 1.21 |
| Irish | 5 | 3.29 | 23 | 2.43 | 0 | 0.00 | 1 | 1.72 | 3053 | 2.63 |
| Mixed | 0 | 0.00 | 0 | 0.00 | 0 | 0.00 | 0 | 0.00 | 13 | 0.01 |
| Other ethnic group | 1 | 0.66 | 5 | 0.53 | 0 | 0.00 | 1 | 1.72 | 1231 | 1.06 |
| Pakistani | 0 | 0.00 | 1 | 0.11 | 0 | 0.00 | 0 | 0.00 | 430 | 0.37 |
| Prefer not to answer | 1 | 0.66 | 4 | 0.42 | 1 | 1.54 | 0 | 0.00 | 400 | 0.34 |
| White | 0 | 0.00 | 3 | 0.32 | 0 | 0.00 | 0 | 0.00 | 115 | 0.10 |
| White and Asian | 0 | 0.00 | 3 | 0.32 | 0 | 0.00 | 0 | 0.00 | 227 | 0.20 |
| White and Black African | 0 | 0.00 | 0 | 0.00 | 1 | 1.54 | 0 | 0.00 | 118 | 0.10 |
| White and Black Caribbean | 0 | 0.00 | 0 | 0.00 | 0 | 0.00 | 0 | 0.00 | 160 | 0.14 |
| **Prospective memory result (first visit)** |  |  |  |  |  |  |  |  |  |  |
| Correct recall on first attempt | 25 | 42.37 | 222 | 71.61 | 20 | 76.92 | 16 | 84.21 | 28604 | 78.35 |
| Correct recall on second attempt | 18 | 30.51 | 60 | 19.35 | 6 | 23.08 | 2 | 10.53 | 6319 | 17.31 |
| Instruction not recalled, either skipped or incorrect | 16 | 27.12 | 28 | 9.03 | 0 | 0.00 | 1 | 5.26 | 1584 | 4.34 |
| **Prospective memory result (second visit)** |  |  |  |  |  |  |  |  |  |  |
| Correct recall on first attempt | 8 | 53.33 | 48 | 75.00 | 6 | 75.00 | 1 | 50.00 | 1171 | 86.48 |
| Correct recall on second attempt | 3 | 20.00 | 16 | 25.00 | 2 | 25.00 | 0 | 0.00 | 153 | 11.30 |
| Instruction not recalled, either skipped or incorrect | 4 | 26.67 | 0 | 0.00 | 0 | 0.00 | 1 | 50.00 | 30 | 2.22 |

**Supplementary Table 3.** Prediction of training and test sets with the multinomial model

| Training set |  |  |  |  |  |
| --- | --- | --- | --- | --- | --- |
|  | Actual diagnosis |  |  |  |  |
| Predicted diagnosis | AD | PD | MND | MG | Control |
| AD | 5 | 1 | 0 | 0 | 3 |
| PD | 13 | 170 | 7 | 5 | 8 |
| MND | 0 | 0 | 0 | 0 | 0 |
| MG | 0 | 0 | 0 | 1 | 0 |
| Control | 8 | 59 | 5 | 5 | 565 |
| Test set |  |  |  |  |  |
|  | Actual diagnosis |  |  |  |  |
| Predicted diagnosis | AD | PD | MND | MG | Control |
| AD | 1 | 0 | 1 | 0 | 0 |
| PD | 8 | 75 | 2 | 3 | 1 |
| MND | 0 | 1 | 0 | 0 | 0 |
| MG | 0 | 0 | 0 | 1 | 0 |
| Control | 2 | 22 | 2 | 1 | 246 |

**Supplementary Table 4.** Leave-one-out cross validation of the multinomial model

| Variable left out | Accuracy |
| --- | --- |
| None | 0.8825 |
| Age when attended assessment centre | 0.8579 |
| Alanine aminotransferase | 0.8825 |
| Albumin | 0.8798 |
| Apolipoprotein A | 0.8770 |
| Calcium | 0.8825 |
| Cholesterol | 0.8798 |
| Cystatin C | 0.8770 |
| Ethnic background | 0.8798 |
| LDL direct | 0.8770 |
| Mean time to correctly identify matches | 0.8770 |
| Microalbumin in urine | 0.8825 |
| Phosphate | 0.8825 |
| Prospective memory result (first visit) | 0.7596 |
| Prospective memory result (second visit) | 0.8825 |
| Sodium in urine | 0.8770 |
| Testosterone | 0.8699 |

**Supplementary Table 5.**
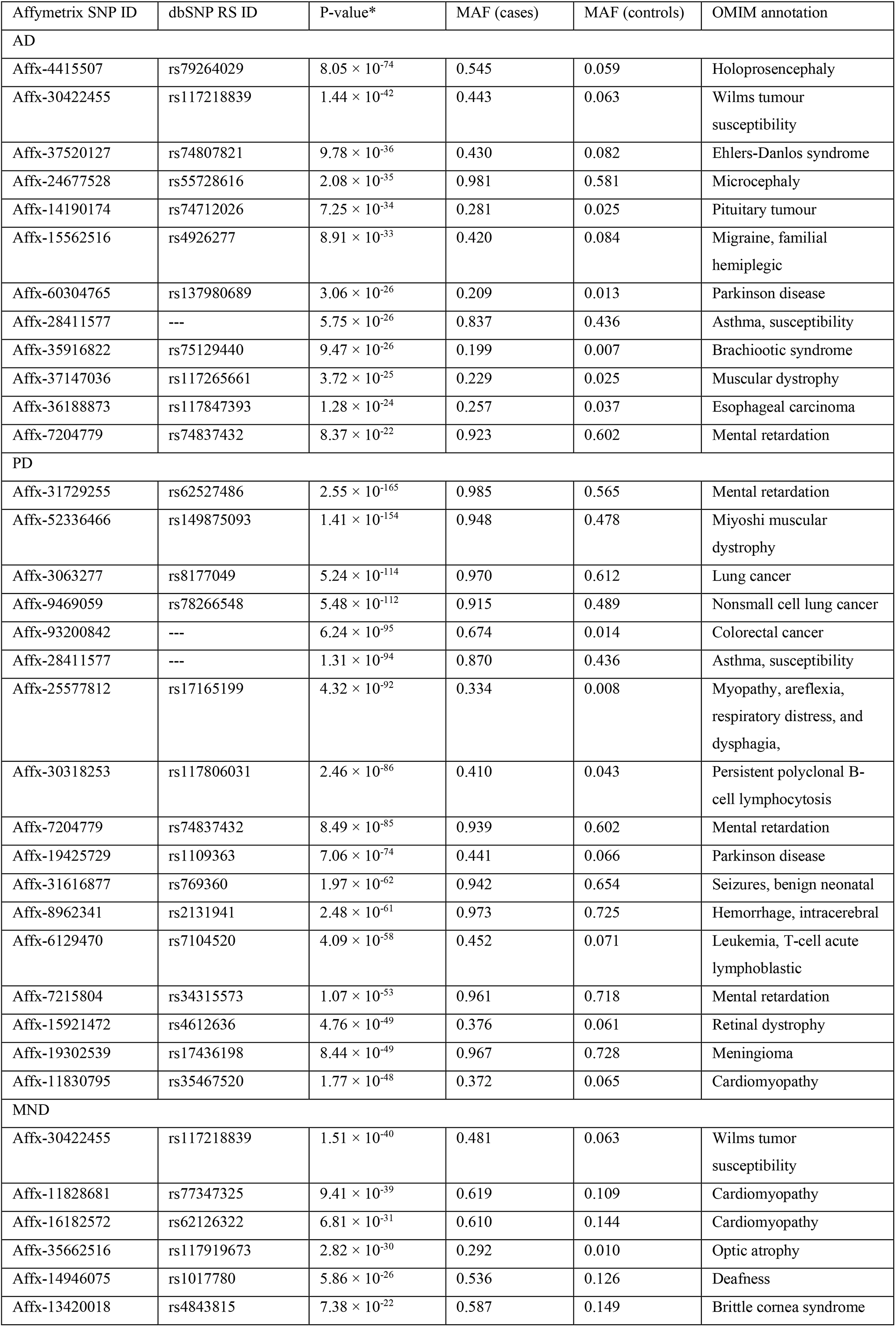
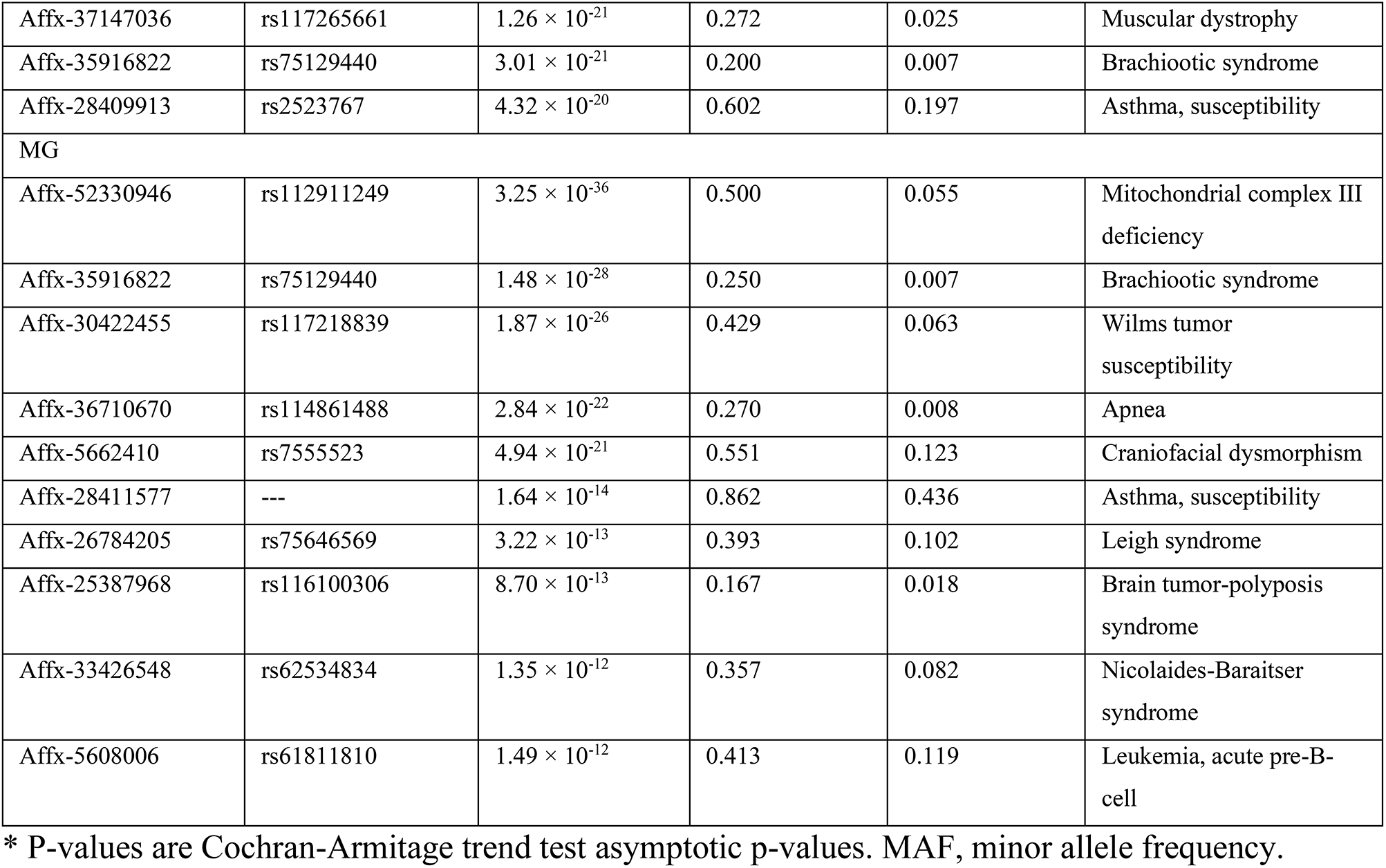
Selected significant SNPs identified in GWAS

